# The frequency of pathogenic variation in the All of Us cohort reveals ancestry-driven disparities

**DOI:** 10.1101/2022.12.19.22283658

**Authors:** Eric Venner, Karynne Patterson, Divya Kalra, Marsha M. Wheeler, Yi-Ju Chen, Sara E. Kalla, Bo Yuan, Jason H. Karnes, Breanna Lee, Kimberly Walker, Josh Smith, Sean Mcgee, Aparna Radhakrishnan, Andrew Haddad, Qiaoyan Wang, Gail Jarvik, Diana Toledo, Anjene Musick, Richard A. Gibbs, the All of Us Research Program Investigators

## Abstract

Disparities in the data that underlies clinical genomic interpretation is an acknowledged problem but there is a paucity of data demonstrating it. The National Institutes of Health’s *All of Us* Research Program aims to collect whole genome sequences, electronic health record (EHR) data, surveys and physical measurements for over a million participants of diverse ancestry and varied access to healthcare resources. We grouped participants by computed genetic ancestry and summarized the frequency of pathogenic variation within these groups. The European subgroup showed the highest rate of pathogenic variation (2.1%), with other ancestry groups ranging from 1.04% (East Asian) to 1.87% (‘Other’). Pathogenic variants were most frequently observed in genes related to Breast/Ovarian Cancer, Hypercholesterolemia or Hemochromatosis. Variant frequencies were consistent with gnomAD and some notable exceptions were resolved using gnomAD subsets. We additionally use this data to enrich sets of participants for specific genetic findings and to calculate penetrance. Differences in the frequency of pathogenic variants observed between ancestral groups generally indicate biases of ascertainment, but some may indicate differences in disease prevalence. These analyses are available on the *All of Us* Researcher Workbench.

## Main

Implementing genomic medicine will require interpreting genomic variation in ‘real world’ clinical populations. At present, lack of diversity in large genomics cohorts is a widely recognized problem^1–4^. Most sequencing studies have focused on European populations^5^ and it is predicted that much of the pathogenic variation present in the general population is specific to an ancestral population^6,7^. Overcoming this gap in diagnostic yield will necessitate collecting diverse genomic data paired with EHR data^8,9^.

To advance precision medicine, the *All of Us* Research Program from the National Institutes of Health (NIH) is generating a unique dataset which includes genetic, electronic health record (EHR) and survey data from a diverse participant cohort^10^. *All of Us* is targeting a cohort size in excess of one million, with a focus on individuals who have been traditionally underserved by biomedical research^10^. Whole genome sequence (WGS) data is generated at one of three *All of Us* Genome Centers located at the Baylor College of Medicine (BCM), the Broad Institute (BI) or the Northwest Genomics Center at the University of Washington (UW). Data are transferred via the *All of Us* Data and Research Center (DRC) at Vanderbilt University to Clinical Validation Laboratories (CVLs) at BCM, UW and Color Genomics, where sequence data are interpreted for health-related reporting. Data are also deposited at the DRC for further processing and sharing with qualified researchers via the *All of Us* Researcher Workbench, a cloud-computing platform. The data generation and results workflow are conducted under an investigational device exemption through the FDA^11^.

A strength of the *All of Us* Research Program is the diversity of the participants, relative to those in previously studied large cohorts (Figure S1). The *All of Us* Researcher Workbench, a cloud computing platform, contains WGS data from 98,622 participants [refer to “Genomic Data in the All of Us Research Program: Advancing Precision Medicine for All”, under review] in a data release called alpha3. Based on genetically predicted ancestry (see Methods), this dataset included 48,351 (49%) participants with predominantly European ancestry, 23,066 (23.4%) participants with predominantly African ancestry, 15,072 (15.3%) American Admixed/Latino participants, 2,116 (2.1%) East Asian participants, 968 (1.0%) South Asian participants, 169 (0.2%) Middle Eastern participants and 8,880 participants which not group unambiguously and were designated as Other. In contrast, the UK Biobank project contains 94% Europeans, the Million Veterans project contains 77% and eMERGE contains 73%.^5^

However, It is currently unknown whether the frequency of pathogenic variants in genes conferring significant health risks in the *All of Us* cohort will differ from those in previously ascertained healthy populations due to the unprecedented diversity of participants enrolled in the program. Identification of such differences will be a powerful reinforcement of the importance of the program’s strategy for recruitment and engagement of participants from underrepresented groups.

To examine the frequency of pathogenic genomic variation in participants we examined data from a set of 73 genes that harbor actionable secondary findings^12^. These genes are associated with diseases, including hereditary breast cancer, hemochromatosis, dislipidemias and cardiomyopathies and represent some of the most well-studied targets for genomic medicine^12^. We annotated each participant with their calculated genetic ancestry and searched for known pathogenic variants with established criteria for pathogenicity, aided by databases of curated variants from previous clinical genomic projects^13,14^, as well as an early “aggregate review” of de-identified *All of Us* data. The preliminary results from analysis of data from more than 98,000 *All of Us* participants showed variability in the rates of pathogenic variants between ancestry groups, prompting further analysis of the source of the differences.

## Results

To understand the rates of previously-known pathogenic variants broken down by predicted ancestry groups in the *All of Us data*, we used the ‘VIP’ database to annotate P/LP variants^15^ present in WGS data in *All of Us* participants across 73 genes with actionable secondary findings^12^. This database contains variants curated by the HGSC-CL variant interpretation group during projects such as eMERGE III^13^ and HeartCare^14^ as well as an initial assessment of deidentified variants from the *All of Us* cohort itself. Figure 1a shows that the European ancestry group has the highest rate of previously known pathogenic variants (2.1%), followed by the “Other” group at 1.87% and the African ancestry group at 1.48%. Using a Chi-square test for independence, the rates of pathogenic variants are significantly different between the African ancestry, European and Admixed American/Latino groups (p < 0.00001), which have sufficient data to perform this test. These differences are significant even when leaving out the gene *HFE*, which has a large known difference in pathogenic rates between ancestry groups. These significant differences in the rates of pathogenic variation could be explained either by a bias in the ascertainment of pathogenic variation in the variant database or by differences in the underlying disease prevalence between ancestry groups.

**Figure 1:**
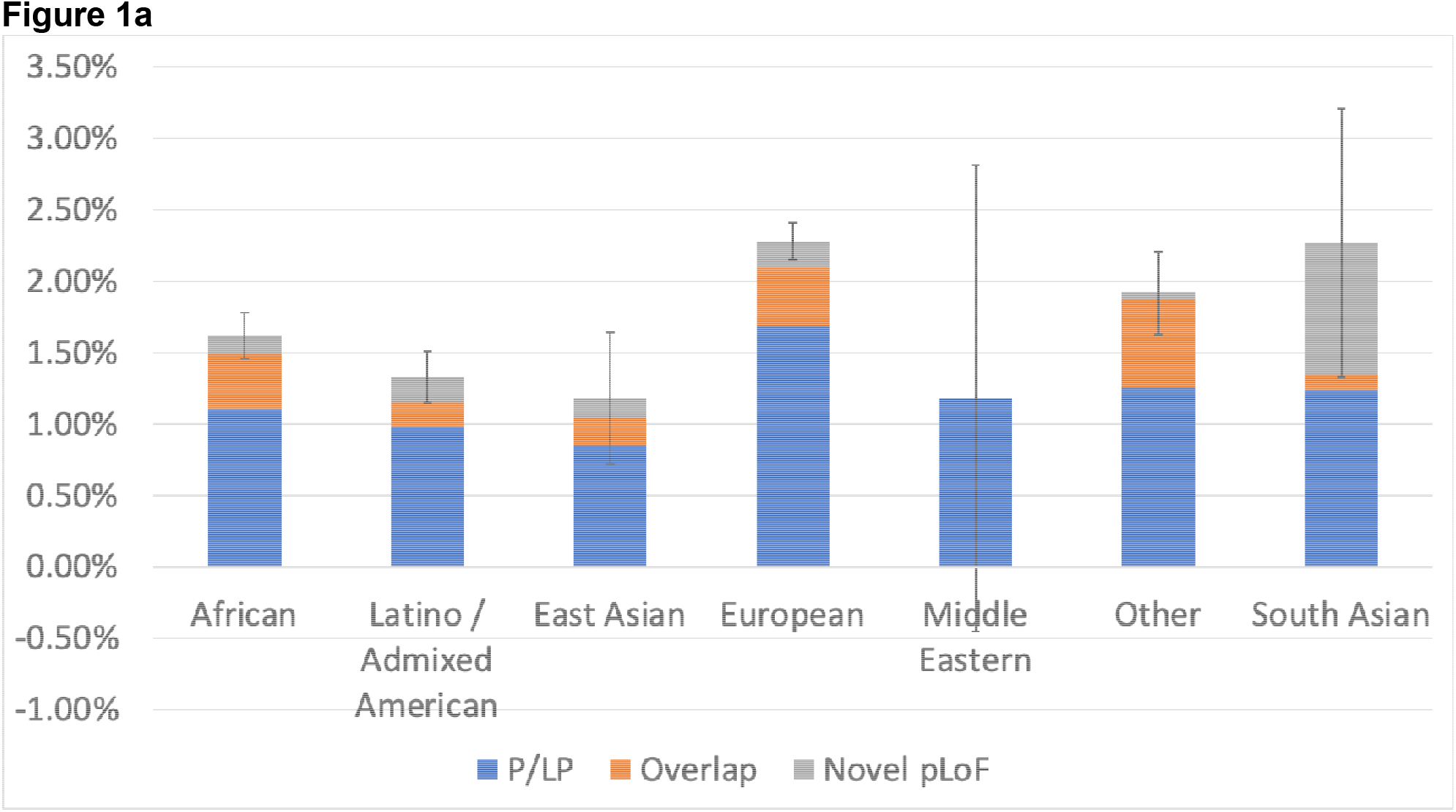
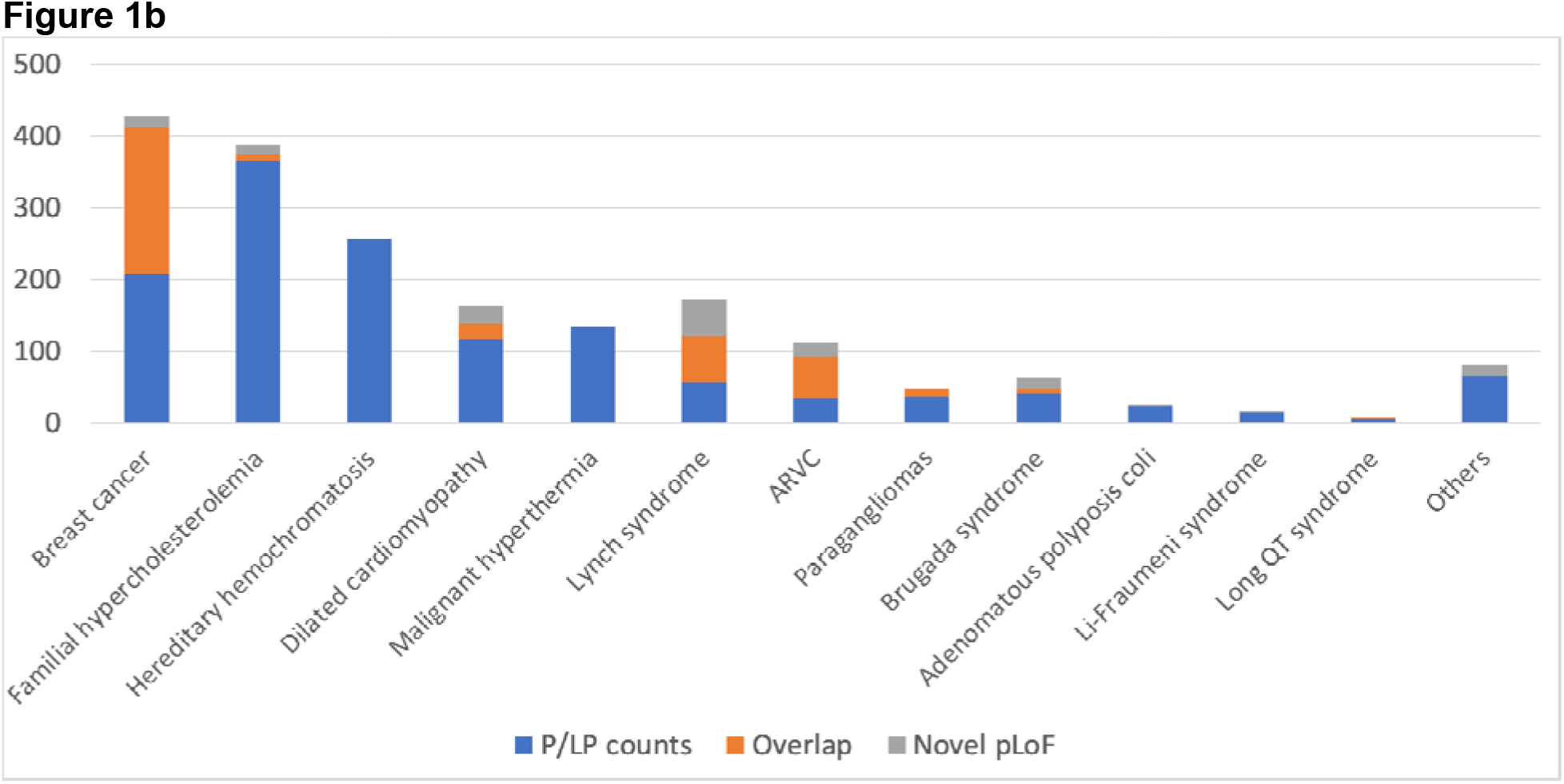
Pathogenic variants by ancestry. Using a database of known pathogenic mutation and annotations for rare, pLoF variants, we searched the beta release of the All of Us cohort for pathogenic variants, using the Researcher Workbench. Figure 1a shows the rates of pathogenic variation, broken down by predicted genetic ancestry groups. Error bars show 95% the confidence intervals for the total set of pathogenic variants (including both VIP P/LP variants and rare pLoF). Figure 1b shows the breakdown of pathogenic variants by disease area.

To build a more comprehensive picture of pathogenic and likely pathogenic variants in these participants, we examined rare (gnomAD popmax allele frequency < 0.001) predicted loss-of-function (pLoF) variants, in a subset of 38 genes (supplementary table S2) in which LoF is a known mechanism of disease, and checked for overlap with known pathogenic variation (Figure 1a, orange and gray bars). We found 236 distinct variants, with 88 frameshifts, 29 splice acceptor variants, 28 splice donor variants, 8 start lost variants, 81 stop gained variants and 2 stop lost variants (Supplemental table 3). These pLoF variants showed significant overlap with the VIP P/LP variants (Figure 1a, orange bar), for example, in the European group, the overlap is 0.41%. Adding in pLoF variants increases the total rates of positive variants, ranging from 2.29% for the European group to 1.18% for the Middle Eastern and East Asian groups, although we note that the smaller groups have large confidence intervals. Interestingly, for rare pLoF variants, the European group no longer has the highest rate of findings with the South Asian (1.03%) and Other (0.65%) having higher rates. The variability of pLoF variants between ancestry groups is lower (pLoF standard deviation of variant rates 0.003 vs 0.004 standard deviation for P/LP variant rates). As interpreting novel LoF variants is simpler and can be automated, these data suggest that ascertainment of pathogenic variants from studies that contain a large number of participants with European ancestry is driving some of the between-group differences.

In order to understand which genes have divergent rates of pathogenic variants between ancestry groups, we normalized pathogenic rates of all genes against the European pathogenic rate and checked for outliers in population proportions using a z-test. Several genes show significant differences from the European rates (Table 1). Interestingly, the genes *HFE* and *PALB2* differences are known^16,17^ but the differences in the gene PKP2 have not been reported previously to our knowledge. Other genes, such as *MYL3* and *APOB* show differences that may indicate altered sources of genetic disease prevalence in some ancestry groups. In the African ancestry subgroup, the *PALB2* and *PKP2* findings were replicated using ClinVar as a source of pathogenic variants instead of the VIP database (supplementary table S4). These gene-level differences present targets for future investigations of health disparities.

**Table 1:** Genes having rates of pathogenicity that differ from the European pathogenic rate.

| Gene | Ancestry group | Ancestry group path. variants | European group path. variants | p value (VIP db) |
| --- | --- | --- | --- | --- |
| HFE (hom) | African | 5 / 23,066 (0.02%) | 235 / 48,351 (0.49%) | 0 |
| APOB | African | 4 / 23,066 (0.017%) | 56 / 48,351 (0.116%) | 0.0001 |
| PKP2 | African | 35 / 23,066 (0.15%) | 19 / 48,351 (0.039%) | 0.000002 |
| PALB2 | African | 32 / 23,066 (0.139%) | 30 / 48,351 (0.062%) | 0.004 |
| HFE (hom) | Admixed American/Latino | 8 / 15072 (0.053%) | 235 / 48,351 (0.49%) | 0 |
| MYL3 | Admixed American/Latino | 7 / 15072 (0.046%) | 0 / 48,351 (0%) | 0.00001 |

In order to understand how these findings compare to other large cohorts, we compared these rates of pathogenic findings to gnomAD. To do so, we first identified pathogenic variation using the VIP database and ClinVar separately. We then annotated these variants with the allele frequencies from the *All of Us* cohort and gnomAD, and summed up the variants in genes to provide gene-level summaries. Under the hypothesis that gnomAD may contain affected individuals, we also made selected comparisons to either the gnomAD non-cancer subset for cancer genes or the non-TopMed subset for genes related to cardiovascular disease. The results (Figure 3) show the relative difference between *All of Us* and gnomAD positive rate, broken down by gene. Overall, we observe high-level concordance between these data sources (Pearson correlation 0.99 between *All of Us* and gnomAD positive rates). However, when comparing gene-level pathogenic rates in ancestry groups other than the European group, some differences are apparent. For example, in the African ancestry group, the incidence of P/LP BRCA2 variants differs between *All of Us* cohort and gnomAD (0.093% vs 0.161%). However, gnomAD is available in different subsets^18^, and when the non-cancer subset is used, the pathogenic rate is very similar to that in the *All of Us* cohort (0.093% vs 0.095%). A similar story is found with LDLR gene in the Admixed American/Latino ancestry group: using a broad comparison between gnomAD and *All of Us*, the rates of pathogenicity are divergent. However, when using the gnomAD non-TopMed subset, the rates of pathogenicity are highly similar, possibly due to dyslipidemia studies in TopMed^19^. Other examples are not as clear. For example, we note divergent pathogenicity rates in the *RYR1* gene between the gnomAD and *All of Us* African ancestry groups, but could not account for them using a gnomAD subset. These results show that the rate of pathogenic variant findings in the *All of Us* are similar to those seen in gnomAD, and the rates become even closer when we account for disease populations within the gnomAD resource.

**Figure 2:**
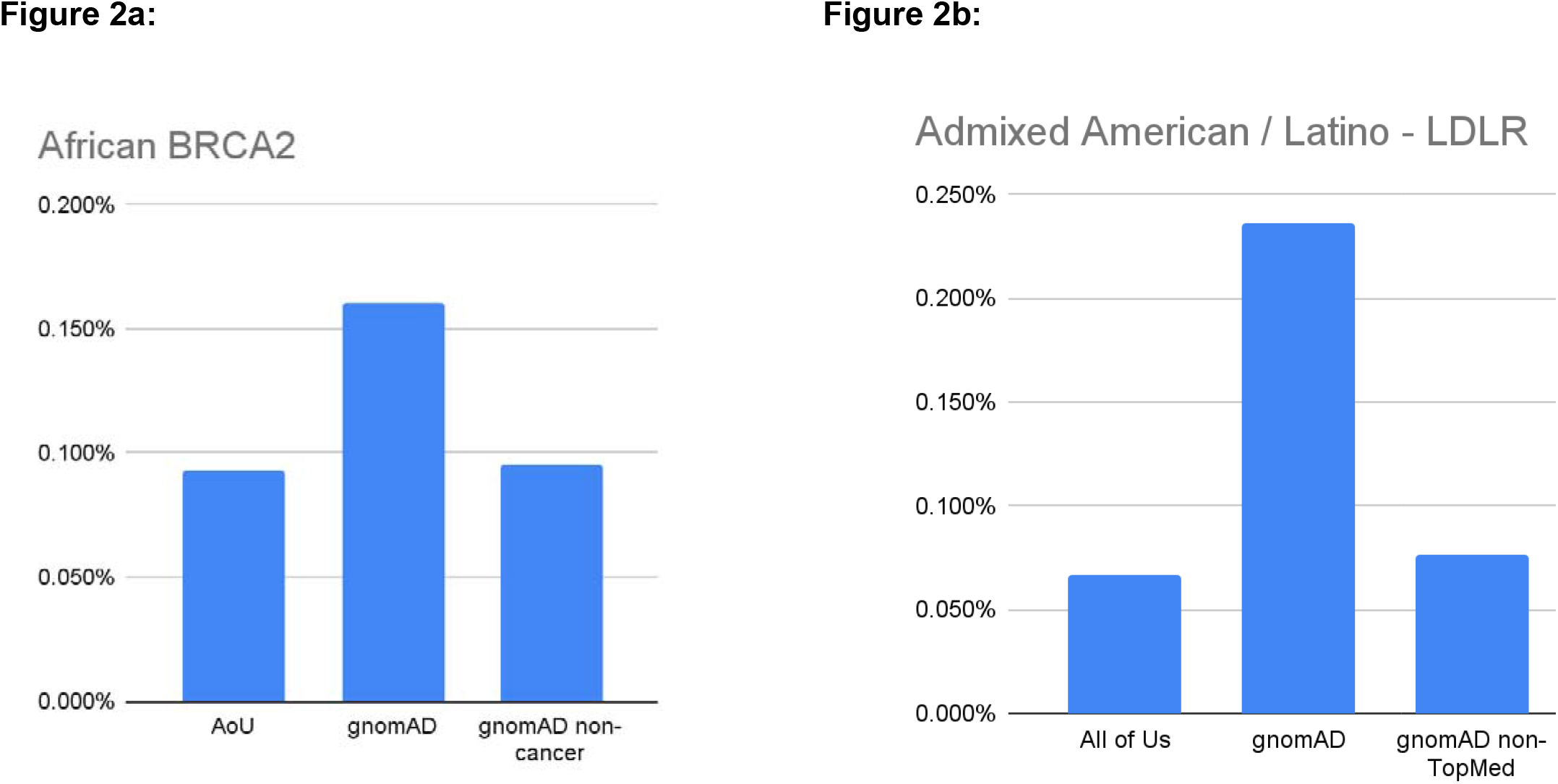
Compar isons to gnomAD subsets. Though there is high level concord ance between the rates of pathogenic variants in All of Us and gnomAD, in some cases there are differences specific to a gene and ancestry group. For example, in participants with African ancestry, the rate of pathogenic variants in *BRCA2* diverges from gnomAD (Figure 2a). However, when the non-cancer subgroup of gnomAD is used, the rates are much more similar. A similar situation is seen in the Admixed American / Latino group with *LDLR*. Using the non-TopMed portion of gnomAD brings the rates much closer.

**Figure 3:**
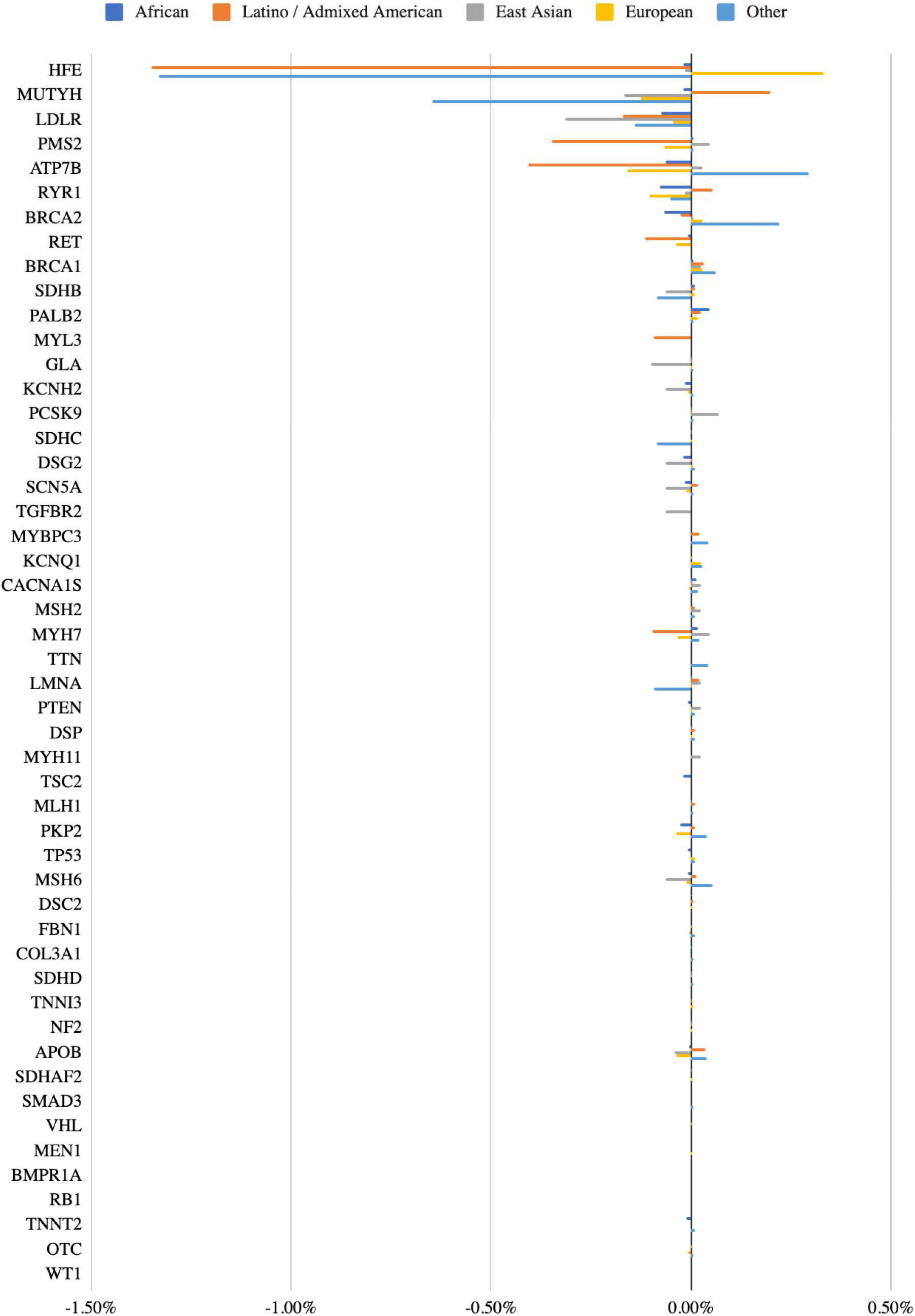
Relative positive rates for *All of U*s vs gnomAD. This figure shows relative frequencies of previously-curated pathogenic or likely pathogenic variants between the *All of Us* cohort and gnomAD, broken down by gene and ancestry group. Overall, there is a high level of concordance between variant frequencies of pathogenic variants; most genes show very small differences relative to gnomAD.

When broken down by disease area, the results show that the predominant health-related findings will be in Breast cancer, familial hypercholesterolemia, Dilated cardiomyopathy and hereditary hemochromatosis (Figure 1b). To further understand how these findings relate to the participant’s available health information, we made use of additional data resources provided in the AoU Researcher Workbench. The *All of Us* Research Workbench provides participant health information in the forms of EHR condition codes as well as survey questions answered by the participants. To begin to understand how this phenotypic information available in the workbench matches with genetic findings, we looked at Breast Cancer as an example. Samples were selected if participants had either “Malignant neoplasm of female breast” or “Malignant tumor of breast” condition codes (SNOMEDCodes 254837009 and 372064008) or if the participant answered “breast cancer” to the question “Has a doctor or health care provider ever told you that you have or had any following cancers?” 8,603 participants fell into this cohort, with 1,653 of those having WGS genetic data thus far. This represents an enrichment for P/LP variants in breast cancer patients (32/1,653, 1.94% vs 414 / 98,622, 0.42%, p < 0.00001), demonstrating the ability to use *All of Us* participant level data to select cohorts enriched for specific genetic factors.

To demonstrate a basic penetrance calculation from this resource, we searched the workbench for participants who had responded to the ‘Personal Medical History’ survey (n=114,454) and had a previously-reported P/LP variant in *BRCA1, BRCA2* or *PALB2* (n=169). Of these, 35 had previously reported that ‘a doctor or healthcare provider had told them they had Breast Cancer, giving a penetrance value of 20.7%. For participants born before 1960, the penetrance increases to 28.8%. The lower penetrance values here compared to those previously reported^20,21^ likely reflect a combination of the incomplete variant interpretation across this cohort, the diverse population suppressing counts of known P/LP variants, and the failure to accurately self-report breast cancer diagnosis.

## Discussion

The diverse cohort collected as part of the *All of Us* will be a rich resource for advancing precision medicine. Here, we have examined the pathogenic variant rate in the beta release of the *All of Us* Researcher Workbench data, finding significant variability between groups of participants with differing ancestry. This variability is likely the result of multiple factors, but ascertainment of pathogenic variants in databases is likely to contribute significantly. Future work will show whether variant interpretation of the *All of Us* diverse cohort will have an impact on this ascertainment bias, but future targeted efforts that aim to perform clinical interpretation of non-European participants could also be necessary.

Ancestry-linked differences in variant pathogenicity found in different genes highlight the importance of the *All of Us’s* diverse cohort. Different ancestry backgrounds likely carry different burdens of risk depending on the frequency of pathogenic haplotypes. Implementing precision medicine will require understanding these varied risk profiles and gathering detailed information on the haplotype structure of the population. Clinical testing is guided to some extent today by ancestry, for example, BRCA2 in the Ashkenazi Jewish population^22^ and HLA-B testing for SJS/TEN in some Asian populations^9,23,24^. As we better understand genetic risk burdens in population groupings we can more precisely target genetic testing and aid interpretation.

Another benefit of this data has been to allow the program to project forward to the return of health-related results, phase of the program. Based on the variants examined in this work, we estimate that 17% of variant interpretations will require manual assessment of literature, which has strong implications for the time required to complete the review of that variant. This in turn has allowed us to better project the resources required to return health related results.

We primarily used an internal database of pathogenic genomic variants in this work (the ‘VIP’ database) with ClinVar^25^ used as a confirmatory resource. Although ClinVar is an essential and widely-used resource, its heterogeneity poses a challenge. For example, the well-known *HFE* NM_000410.4:c.845G>A variant, which we previously reported clinically for the eMERGE III^13^ project when in the homozygous state, is associated with hemochromatosis. However, at the time of publication, this variant is listed as “conflicting” in ClinVar and so was excluded under our simple ClinVar filtering scheme (see Methods). Many other variants likely fall into a similar scenario. It would be possible to fine-tune filters for ClinVar variants and reanalyze this data, which may provide a more complete picture of known pathogenic variation, but there are large effects on sensitivity and specificity that arise due to this tuning that would need to be understood. This level of curation was beyond the scope of the current study.

As a baseline for our expectations of the frequency of pathogenic variants we have compared to gnomAD^26^. At a high-level, we found strong concordance between these cohorts, although there are some outliers when individual gene-level frequencies are examined. Some differences are likely due to the cohorts applying slightly different ancestry definitions. For example, the precise definition of the “Other” group is likely to be different, and the gnomAD counts do not include the Middle Eastern and South Asian groups. Using local ancestry (i.e. assigning ancestry not at the sample-level but at the haplotype level) would help resolve this issue.

The *All of Us* Researcher Workbench enabled this first assessment of pathogenic variants within the *All of Us* cohort, and the diversity of that population has allowed us to begin to detect different frequencies in pathogenic variation between several of the ancestry groups. More work in this area will further reveal groups of participants who carry under-studied pathogenic variation and allow us to target precision medicine efforts at those disparities.

## Online Methods

### *All of Us* Demonstration Projects

The goals, recruitment methods and sites, and scientific rationale for *All of Us* have been described previously^27^. Demonstration projects were designed to describe the cohort, replicate previous findings for validation, and avoid novel discovery in line with the program values to ensure equal access by researchers to the data^28^.The work described here was proposed by Consortium members, reviewed and overseen by the program’s Science Committee, and confirmed as meeting criteria for non-human subjects research by the *All of Us* Institutional Review Board. The initial release of data and tools used in this work was published recently^28^.

### *All of Us* Research Hub

This work was performed on data collected by the previously described *All of Us* Research Program using the *All of Us* Researcher Workbench, a cloud-based platform where approved researchers can access and analyze *All of Us* data. The *All of Us* data currently includes surveys, EHRs, and physical measurements (PM). The details of the surveys are available in the Survey Explorer found in the Research Hub, a website designed to support researchers^3^. Each survey includes branching logic and all questions are optional and may be skipped by the participant. PM recorded at enrollment include systolic and diastolic blood pressure, height, weight, heart rate, waist and hip measurement, wheelchair use, and current pregnancy status. EHR data was linked for those consented participants. All three datatypes (survey, PM, and EHR) are mapped to the Observational Medicines Outcomes Partnership (OMOP) common data model v 5.2 maintained by the Observational Health Data Sciences and Informatics (OHDSI) collaborative. To protect participant privacy, a series of data transformations were applied. These included data suppression of codes with a high risk of identification such as military status; generalization of categories, including age, sex at birth, gender identity, sexual orientation, and race; and date shifting by a random (less than one year) number of days, implemented consistently across each participant record. Documentation on privacy implementation and creation of the Curated Data Repository (CDR) is available in the *All of Us* Registered Tier CDR Data Dictionary^4^. The Researcher Workbench currently offers tools with a user interface (UI) built for selecting groups of participants (Cohort Builder), creating datasets for analysis (Dataset Builder), and Workspaces with Jupyter Notebooks (Notebooks) to analyze data. The Notebooks enable use of saved datasets and direct query using R and Python 3 programming languages.

#### Samples/Dataset

Aggregate data for this study was generated from *All of Us* participant data (N=98,622) using the *All of Us* Researcher Workbench cloud computing platform. We accessed variant data (single nucleotide variants (SNVs) and indels) from WGS in the alpha3 data release provided by the *All of Us* DRC. Details regarding *All of Us* DRC’s genomic pipelines can be found here [refer to “Genomic Data in the All of Us Research Program: Advancing Precision Medicine for All”, under review]. *All of Us* variant data were generated using the GRCh38 human reference build and made available on the pre-production version of the *All of Us* Researcher Workbench using the Hail framework^29^.

For our analyses, we subsetted the whole genome variants Hail matrix table to coding regions for the 73 genes listed in the ACMG SF v3.0 list [PMID: 34012068]. In these 73 genes, we also included regions 2000bp upstream and 1000bp downstream of the coding regions to ensure we do not exclude any pathogenic variants outside of the coding regions. Variants were filtered on genotype quality (GQ > 20) and variant pathogenicity (using ‘Path’ or ‘LPath’ values in the Vip_variant_interpretation field from the VIP database). These variants were then grouped by predicted genetic ancestry and genes to build a contingency table, with heterozygous variant counts across most genes. For the autosomal recessive genes *MUTYH, ATP7B* and *KCNQ1*, only homozygous variants were counted. In *HFE*, only the *NM_000410*.*4:c*.*845G>A* variant in the homozygous state was considered. We annotated *All of Us* participants with predicted genetic ancestry provided by the *All of Us* DRC.

For predicting genetic ancestry, briefly, a total of 56,671 high quality variant sites from chromosomes 20 and 21 were selected in both the training dataset (Human Genome Diversity Project + 1000 Genomes) and the WGS dataset to generate the first sixteen principal components (PCs) using Hail. These PCs were then used as the features for a random forest classifier. Samples were assigned an ancestry group if the classification probability exceeded 90%. The ancestry super-populations correspond to the ancestry definitions used within gnomAD^26^, the Human Genome Diversity Project^30,31^, and 1000 Genomes^6^. These include African/African American (afr), American Admixed/Latino (amr), East Asian (eas), European (eur), Middle Eastern (mid), South Asian (sas), and Other (oth; not unambiguously clustering with super-population in the principal component analysis (PCA))^30^.

#### Annotation of known pathogenic variants

For annotation of known pathogenic or likely pathogenic (P/LP) variants, we used a database of variants (the ‘VIP’ database) collected during clinical reporting activities at the Human Genome Sequencing Center-Clinical Laboratory (HGSC-CL). An advantage of this ‘VIP’ database is that the variant curations have been performed by the HGSC-CL’s Clinical Variant Interpretation team, using a consistent set of criteria, whereas resources like ClinVar can contain conflicting assertions of variant pathogenicity. The ‘VIP’ database used in this study contains 59,383 genomic variants, including pathogenic, likely pathogenic, variants of uncertain significance (VUS), benign, likely benign and risk alleles, across 583 genes (Supplemental file vip_gene_count.xslx). The majority were assessed during prior HGSC-CL projects, including the NIH eMERGE III program^13^ and HeartCare^14^, a local cardiovascular risk assessment project. All variants were interpreted following Association for Molecular Pathology/American College of Medical Genetics and Genomics (AMP/ACMG) guidelines^32^.

To detect predicted loss of function (pLoF) variants, we additionally annotated aggregate variant data using Variant Effect Predictor (VEP) with the LOFTEE plugin^26^. pLoF variants were filtered, retaining only those with ‘high confidence’. The following variant effects were treated as loss of function: “frameshift_variant”, “stop_gained”, “stop_lost”, “splice_acceptor_variant, “splice_donor_variant”, and “start_lost” when they were seen in the “vep.most_severe_consequence” field. Other VEP “HIGH” impact effects such as “transcript_ablation”^33^) were not present in the dataset.

#### Statistical analyses

The comparison of pathogenic variant counts between predicted genetic ancestry groups, in the *All of Us* dataset, was done with a Chi-Square test for independence. To meet the Chi-square requirement that all entries have at least a count of 5, the less-represented ancestries (East Asian, Middle Eastern, South Asian and Other) and genes were aggregated into a single column and row in the contingency table, respectively (Supplemental table 1). To detect genes with outlying rates of P/LP variants, we compared proportions to the European rate under the null hypothesis that the proportions are equal. We used a Z-test statistic:

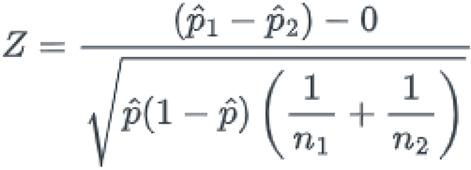

Where p^ is the overall proportion:

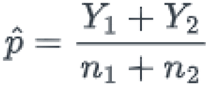

Y1 and Y2 are the group (i.e. participants with primarily European ancestry vs participants with primarily African ancestry) variant counts, n1 and n2 are the group totals and p1^ and p2^ are the group proportions. To translate the Z-score to a p-value we assumed a two-tailed normal distribution.

ClinVar comparisons used variant_summary.txt from Jan, 22, 2022, downloaded from the ClinVar downloads site^34^. We preprocessed this file by 1) filtering out GRCh37 data from the downloaded file and using GRCh38 entries only and 2) filtering the variants 2 stars and above where the ClinicalSignificance field is “Likely pathogenic” or “Likely pathogenic, risk factor” or “Pathogenic” or “Pathogenic/Likely pathogenic” or “Pathogenic/Likely pathogenic, risk factor” or “Pathogenic, risk factor” and the ReviewStatus field is “criteria provided, multiple submitters, no conflicts”, and the LastEvaluated date is on or after Jan 1, 2016. We included three and four star pathogenic variants (“reviewed by expert panel” and “practice guideline” respectively) regardless of the last evaluated time.

GnomAD data used gnomAD v2.1.1 liftover data set from the download site^35^. For ClinVar, we chose entries from 2016 or later and having two or more stars. We joined the gnomAD and ClinVar dataset by using a variant’s chromosome-position-ref-alt combination as a primary key. We then calculated the pathogenic and likely pathogenic (P/LP) ratio in each gene by adding up the alternate allele count in each gene as the numerator and used the maximum of total alleles in each gene as the denominator. We calculated P/LP ratio/frequency in each gene in the dataset, building a contingency table that mirrored the format derived from that from the *All of Us* variant data.

## Data Availability

All data produced are available in the All of Us researcher workbench.

https://www.researchallofus.org/data-tools/workbench/

## Acknowledgements

The All of Us Research Program is supported by the National Institutes of Health, Office of the Director: Regional Medical Centers: 1 OT2 OD026549; 1 OT2 OD026554; 1 OT2 OD026557; 1 OT2 OD026556; 1 OT2 OD026550; 1 OT2 OD 026552; 1 OT2 OD026553; 1 OT2 OD026548; 1 OT2 OD026551; 1 OT2 OD026555; IAA #: AOD 16037; Federally Qualified Health Centers: HHSN 263201600085U; Data and Research Center: 5 U2C OD023196; Biobank: 1 U24 OD023121; The Participant Center: U24 OD023176; Participant Technology Systems Center: 1 U24 OD023163; Communications and Engagement: 3 OT2 OD023205; 3 OT2 OD023206; and Community Partners: 1 OT2 OD025277; 3 OT2 OD025315; 1 OT2 OD025337; 1 OT2 OD025276. In addition, the All of Us Research Program would not be possible without the partnership of its participants.

We thank our colleagues, Kelsey Mayo, Ashley Able, Ashley Green, Andrea Ramirez, and Sokny Lim for providing their support and input throughout the demonstration project lifecycle. We thank Dr. Jun Qian and Dr. Lina Sulieman for providing input on the project’s code review. We thank Lee Lichtenstein and Jennifer Zhang for providing the data artifacts used for the project. We thank the DRC’s Research Support team for their help during implementation. We also thank the *All of Us* Science Committee and *All of Us* Steering Committee for their efforts evaluating and finalizing the approved demonstration projects. The *All of Us* Research Program would not be possible without the partnership of contributions made by its participants. See the <u>supplementary information</u> for a roster of past and present *All of Us* principle investigators. To learn more about the *All of Us* Research Program’s research data repository, please visit https://www.researchallofus.org/.

## Data availability

All data reside in the *All of Us* Researcher Workbench in the ‘Demo - Assessment of pathogenic variants across the All of Us Research Program’ project on the AoU Research Workbench. The VIP database is available on gitlab: https://gitlab.com/bcm-hgsc/neptune.

## Supplementary Tables

**Table S1:**

| Gene | afr | amr | eur | eas, mid, sas, oth | total |
| --- | --- | --- | --- | --- | --- |
| HFE (hom) | 5 | 8 | 235 | 8 | 256 |
| BRCA2 | 43 | 27 | 110 | 44 | 224 |
| LDLR | 71 | 20 | 102 | 27 | 220 |
| RYR1 | 17 | 16 | 76 | 11 | 120 |
| BRCA1 | 24 | 10 | 71 | 14 | 119 |
| MYBPC3 | 16 | 6 | 73 | 8 | 103 |
| APOB | 4 | 11 | 56 | 8 | 79 |
| PALB2 | 32 | 8 | 30 | 1 | 71 |
| PKP2 | 35 | 3 | 19 | 7 | 64 |
| MYH7 | 14 | 6 | 30 | 6 | 56 |
| PMS2 | 9 | 2 | 41 | 3 | 55 |
| others | 72 | 58 | 174 | 66 | 370 |
| total | 342 | 175 | 1017 | 203 | 1737 |

**Table S2:** pLoF genes.

| Gene | Transcript | LoF Mechanism? | Dosage Score |
| --- | --- | --- | --- |
| ACTA2 | NM_001613.3 | NO | 0 |
| *ACTC1* | NM_005159.4 | NO | 1 |
| APOB | NM_000384.2 | NO | 0 |
| CACNA1S | NM_000069.2 | NO | 0 |
| *DSC2* | NM_024422.3 | NO | 2 |
| *DSG2* | NM_001943.4 | NO | 1 |
| MYH11 | NM_001040113.1 | NO | 0 |
| MYH7 | NM_000257.3 | NO | 0 |
| *MYL2* | NM_000432.3 | NO | 30 |
| MYL3 | NM_000258.2 | NO | 0 |
| PCSK9 | NM_174936.3 | NO | 40 |
| *PRKAG2* | NM_016203.3 | NO | 0 |
| *RET* | NM_020975.5 | NO | 3 |
| *RYR1* | NM_000540.2 | NO | 0 |
| *RYR2* | NM_001035.2 | NO | 0 |
| *SDHAF2* | NM_017841.2 | NO | 2 |
| TGFBR1 | NM_004612.3 | NO | 2 |
| TGFBR2 | NM_003242.5 | NO | 2 |
| *TMEM43* | NM_024334.2 | NO | 0 |
| TNNI3 | NM_000363.4 | NO | 1 |
| TNNT2 | NM_001001430.1 | NO | 0 |
| TPM1 | NM_001018005.1 | NO | 0 |
| *APC* | NM_000038.5 | YES | 3 |
| ATP7B | NM_000053.3 | YES | 30 |
| *BMPR1A* | NM_004329.2 | YES | 3 |
| BRCA1 | NM_007294.3 | YES | 3 |
| BRCA2 | NM_000059.3 | YES | 3 |
| *COL3A1* | NM_000090.3 | YES | 3 |
| *DSP* | NM_004415.2 | YES | 1 |
| *FBN1* | NM_000138.4 | YES | 3 |
| *GLA* | NM_000169.2 | YES | 3 |
| *KCNH2* | NM_000238.3 | YES | 3 |
| *KCNQ1* | NM_000218.2 | YES | 3 |
| *LDLR* | NM_000527.4 | YES | 3 |
| *LMNA* | NM_005572.3 | YES | 2 |
| *LMNA* | NM_170707.2 | YES | 2 |
| *MEN1* | NM_130799.2 | YES | 3 |
| *MLH1* | NM_000249.3 | YES | 3 |
| *MSH2* | NM_000251.2 | YES | 3 |
| *MSH6* | NM_000179.2 | YES | 3 |
| *MUTYH* | NM_001128425.1 | YES | 30 |
| *MYBPC3* | NM_000256.3 | YES | 3 |
| *NF2* | NM_000268.3 | YES | 3 |
| *OTC* | NM_000531.5 | YES | 3 |
| *PKP2* | NM_004572.3 | YES | 3 |
| *PMS2* | NM_000535.6 | YES | 3 |
| *PTEN* | NM_000314.6 | YES | 3 |
| *RB1* | NM_000321.2 | YES | 3 |
| *SCN5A* | NM_198056.2 | YES | 2 |
| *SDHB* | NM_003000.2 | YES | 3 |
| *SDHC* | NM_003001.3 | YES | 3 |
| *SDHD* | NM_003002.3 | YES | 3 |
| *SMAD3* | NM_005902.3 | YES | 3 |
| *SMAD4* | NM_005359.5 | YES | 3 |
| *STK11* | NM_000455.4 | YES | 3 |
| *TP53* | NM_000546.5 | YES | 3 |
| *TSC1* | NM_000368.4 | YES | 3 |
| *TSC2* | NM_000548.3 | YES | 3 |
| *VHL* | NM_000551.3 | YES | 3 |
| *WT1* | NM_024426.5 | YES | 3 |

**Table S3:**

| Gene\Consequence | frameshift_variant | splice acceptor variant | splice donor variant | start lost | stop gained | stop lost |
| --- | --- | --- | --- | --- | --- | --- |
| APC | 0 | 0 | 1 | 0 | 0 | 0 |
| ATP7B | 14 | 3 | 4 | 1 | 5 | 0 |
| BRCA1 | 9 | 4 | 3 | 1 | 8 | 1 |
| BRCA2 | 27 | 3 | 1 | 0 | 9 | 0 |
| DSP | 2 | 0 | 1 | 0 | 4 | 0 |
| KCNH2 | 1 | 0 | 1 | 0 | 1 | 0 |
| KCNQ1 | 2 | 0 | 1 | 0 | 1 | 0 |
| LDLR | 2 | 2 | 3 | 0 | 3 | 0 |
| LMNA | 0 | 0 | 1 | 0 | 1 | 0 |
| MLH1 | 0 | 0 | 0 | 0 | 3 | 0 |
| MSH2 | 2 | 0 | 1 | 0 | 2 | 0 |
| MSH6 | 12 | 2 | 3 | 0 | 8 | 0 |
| MUTYH | 4 | 3 | 2 | 2 | 12 | 1 |
| MYBPC3 | 1 | 1 | 1 | 1 | 3 | 0 |
| PKP2 | 8 | 3 | 1 | 0 | 3 | 0 |
| PMS2 | 3 | 1 | 1 | 1 | 5 | 0 |
| SCN5A | 0 | 1 | 0 | 0 | 4 | 0 |
| SDHB | 0 | 0 | 0 | 1 | 2 | 0 |
| SDHC | 0 | 1 | 0 | 0 | 1 | 0 |
| TP53 | 0 | 1 | 0 | 0 | 0 | 0 |
| TSC1 | 0 | 1 | 0 | 0 | 1 | 0 |
| TSC2 | 0 | 3 | 2 | 0 | 2 | 0 |
| VHL | 0 | 0 | 0 | 1 | 1 | 0 |
| WT1 | 1 | 0 | 1 | 0 | 2 | 0 |

**Table S4:**

| Gene | Ancestry group | Ancestry group path. variants | European group path. variants | p value |
| --- | --- | --- | --- | --- |
| PKP2 | African | 28/23066 (0.12%) | 15/48351 (0.031%) | 0.00002 |
| PALB2 | African | 36 / 23,066 (0.15%) | 30 / 48,351 (0.062%) | 0.0005 |

## Supplementary Figures

**Figure S1:**
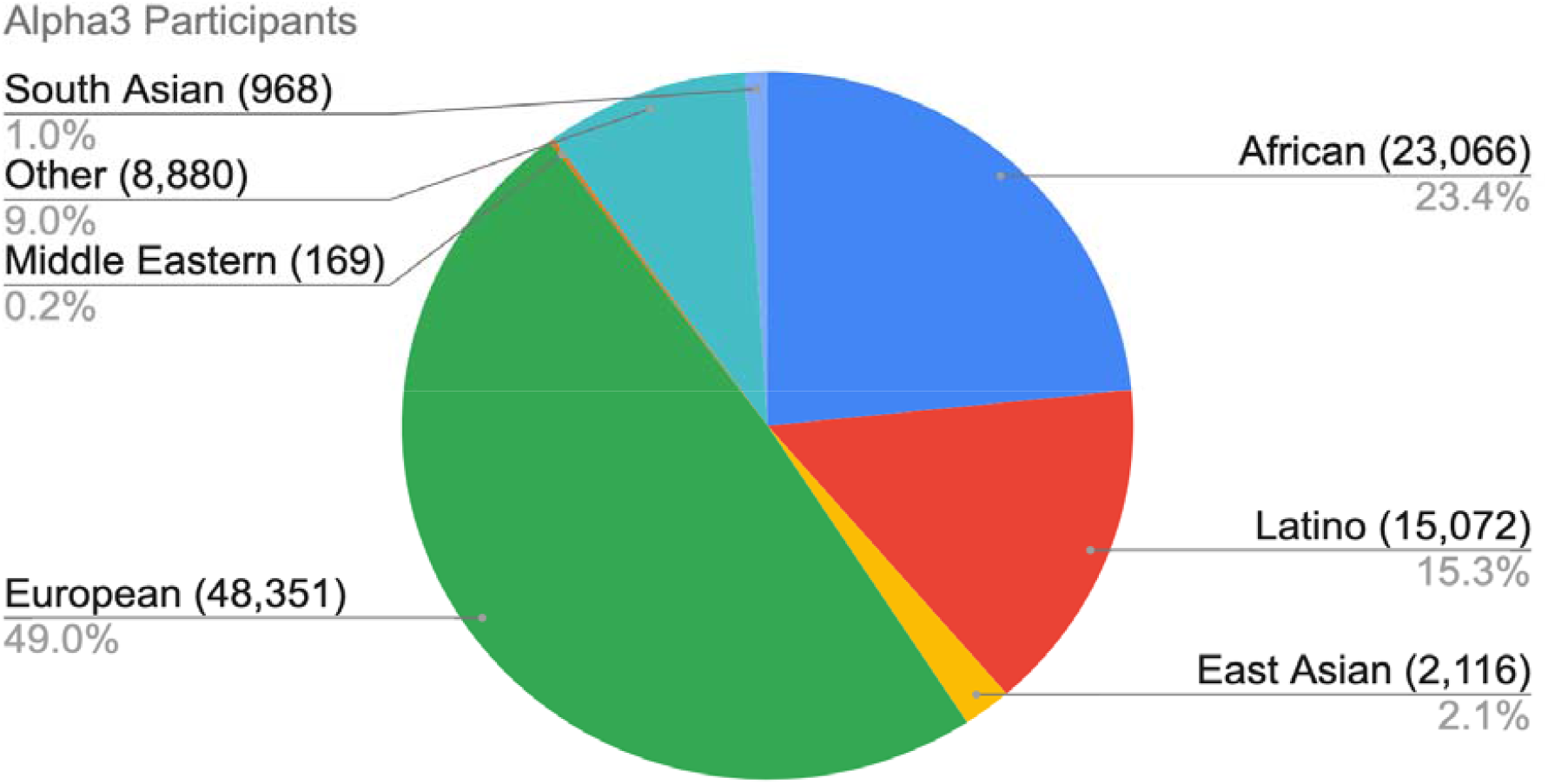

